# Changing patterns of heart failure in China from 1990 to 2021: a secondary analysis of the Global Burden of Disease study 2021

**DOI:** 10.1101/2024.10.24.24316080

**Authors:** Qiwen Yang, Rui Zhuang, Diyang Lyu, Donghua Xue, Chaofeng Niu, Yujie Shi, Meng Li, Lijing Zhang

## Abstract

**Background:** Heart failure is a leading public health issue in China, with a steadily increasing burden. This study aims to assess the changing patterns of heart failure in China from 1990 to 2021, providing evidence for informed healthcare strategies.

**Methods:** Data on prevalence, years lived with disability (YLDs), and their corresponding 95% uncertainty intervals (UI) were obtained from the Global Burden of Disease (GBD) Study 2021. The joinpoint regression model was used to identify both overall and localized trends of heart failure burden, and the age-period-cohort model served to analyze the contributions of age, period, and birth cohort separately. We further utilized the autoregressive integrated moving average (ARIMA) model to predict future trends of heart failure in the next 10 years.

**Results:** In 2021, 13099727 (95% UI, 11320895 to 15376467) individuals lived with heart failure and this illness accounted for 1290810 (95% UI, 865894 to 1775731) YLDs in China. The burden of heart failure is more pronounced in males and the elderly, and ischemic heart disease has become the leading cause since 2002. The age-standardized rates of prevalence and YLDs increased at average annual percentage changes of 0.23% (95% CI, 0.20 to 0.26) and 0.25% (95% CI, 0.23 to 0.27) respectively. The curve of local drift showed a downward trend with age. Both the period and cohort rate ratios have increased significantly over the last 30 years. By 2031, the age-standardized rates of prevalence will decrease to 678.69 (95% CI, 640.75 to 716.63), while the age-standardized rates of YLDs will increase to 69.19 (95% CI, 66.95 to 71.43).

**Conclusions:** The burden of heart failure in China remains concerning. The implementation of comprehensive strategies should be taken into consideration, including strengthening primary healthcare system, enhancing public awareness, and promoting cardiac rehabilitation.

**Clinical Perspective:** *What Is New?:*

- This is the first study of the Global Burden of Disease (GBD) 2021 that comprehensively analyzes the burden of heart failure in China over the past 30 years.

*What Are the Clinical Implications?:*

- The burden of heart failure in China remains concerning.
- Comprehensive strategies prioritizing primary healthcare system, public awareness, and cardiac rehabilitation could be effective to mitigate the burden of heart failure.

## Introduction

Heart failure (HF) is a complex syndrome often identified by characteristic symptoms caused by structural or functional defects in ventricular filling or ejection of blood.^1, 2^ Approximately 55.5 million people worldwide suffer from HF,^3^ which poses notable clinical and public health challenges, including impaired quality of life,^2, 4^ high rates of hospitalization,^5, 6^ poor prognosis,^7, 8^ and huge economic burden.^9, 10^ Therefore, strategies for the prevention and management are important to minimize the burden of HF.

The Global Burden of Diseases, Injuries, and Risk Factors (GBD) study aims to quantify the comparative magnitude of health loss, which provides an opportunity to understand in a given place, time, and age-sex group what are the most important contributors to health loss.^11^ Two recent secondary analyses based on GBD 2019 illustrated the burden, trends, and regional variations in global HF from different perspectives.^12, 13^ However, due to the scope defined by their primary objectives, they were unable to provide detailed country- or region-specific analyses. In the past 30 years, China has made remarkable progress in the Healthcare Quality and Access (HAQ) Index, ranking the first among middle-income countries.^14^ Still, China is struggling with steadily increasing prevalence of HF and high mortality rate.^15, 16^ With China being the world’s most populous country, it contributes to nearly one-third of all HF cases globally, highlighting its substantial burden on both national and international levels.^13^ As such, there is an urgent need to thoroughly analyze the GBD data in China and implement targeted measures to reduce HF burden. Furthermore, recently released GBD 2021 updated new etiologies and methodologies, which led to a significant variation in the data.^3, 17^ For example, regarding China’s HF burden in 2019, GBD 2019 reported an age-standardized prevalence of 1032.84 per 100000 population,^12^ compared to 690.94 per 100000 population as reported in GBD 2021 (https://vizhub.healthdata.org/gbd-results/). Considering such a large discrepancy could skew the understanding of HF in China, it is crucial to use the most recent data.

This first-to-date study provides a comprehensive analysis of HF data in China using GBD 2021. We (1) quantified the burden and trends of HF over the past 30 years by sex, age, and cause; (2) assessed localized trend changes; (3) disentangled the complex effects of age, period, and birth cohort on HF burden; and (4) predicted the HF burden in the next decade. Our findings are expected to provide essential evidence for further healthcare strategies that can mitigate HF.

## Methods

### Data source

The GBD 2021 synthesized a vast array of data input sources and estimated the incidence, prevalence, years lived with disability (YLDs), years of life lost (YLLs), disability-adjusted life-years (DALYs), and healthy life expectancy (HALE) for 371 diseases and injuries across 204 countries and territories, as well as 811 subnational areas, covering the period from 1990 to 2021.^3^ All estimates for China and globally were obtained from the GBD Results Tool (https://vizhub.healthdata.org/gbd-results/).

A total of seven national or regional epidemiological studies were utilized as data sources for HF in China in GBD 2021. Detailed information on these data sources and their characteristics is accessible through the GBD 2021 Sources Tool (https://ghdx.healthdata.org/gbd-2021/sources). The modelling software MR-BRT (meta-regression—Bayesian, regularized, trimmed) was employed to adjust systematic bias between definitions of HF. And overall prevalence of HF was estimated in DisMod-MR 2.1 (Disease Modelling Meta-Regression, version 2.1) based on the input data. More information on data processing and modeling methodologies can be found elsewhere.^3^

This study followed the Guidelines for Accurate and Transparent Health Estimation Reporting (GATHER) to ensure the transparency and reliability of results, and the GATHER checklist is presented in **Supplemental Table S1**.

### Definition and causal attribution

HF, as a level 1 impairment in GBD 2021, is diagnosed using structured criteria, including the Framingham or European Society of Cardiology (ESC) criteria.^3^ Briefly, the Framingham criteria need to be satisfied with at least two major criteria or one major and two minor criteria,^18^ whereas the ESC criteria require typical signs and symptoms caused by a structural and/or functional cardiac abnormality, resulting in a reduced cardiac output and/or elevated intracardiac pressures at rest or during stress.^19^ HF is further divided into four subgroups based on clinical symptoms: treated, mild, moderate and severe HF.^3^

HF can be attributed to 20 level 3 causes, as defined in a previous study,^3^ including: atrial fibrillation and flutter; cardiomyopathy and myocarditis; chagas disease; chronic kidney disease (CKD); chronic obstructive pulmonary disease (COPD); cirrhosis and other chronic liver diseases; congenital birth defects; drug use disorders; endocarditis; endocrine, metabolic, blood, and immune disorders; hemoglobinopathies and hemolytic anemias; hypertensive heart disease; interstitial lung disease and pulmonary sarcoidosis; ischemic heart disease; non-rheumatic valvular heart disease; other cardiovascular and circulatory diseases; pneumoconiosis; pulmonary arterial hypertension; rheumatic heart disease; and stroke.

### Statistical analysis

We used prevalence and YLDs to quantify the burden of HF in China during 1990-2021 by sex, age, and cause. Age-standardized rates (ASRs) of prevalence and YLDs were also used to avoid the influences of population size and age composition. YLDs were measured by taking the prevalence multiplied by the disability weight for HF.^3^ 95% uncertainty interval (UI) was generated for all final estimates as the 2.5th and 97.5th percentiles values of 500 draws.^3^

Joinpoint software (Version 5.2.0, https://surveillance.cancer.gov/joinpoint/) was utilized to further assess trends in HF over the past 30 years. The joinpoint regression, a segmented linear model, is designed to identify both overall and localized trends in data changes.^20^ Annual percentage changes (APC) reflect shifts in the slope within each time segment, while average annual percentage changes (AAPC), calculated by weighting segment lengths, provide an overall trend for the entire period. APC and AAPC are considered stable if their 95% confidence interval (CI) included 0. A *P*-value of less than 0.05 indicates a significant increasing or decreasing trend.

The age-period-cohort (APC) model helps to assess the contribution of age, period, and cohort to the burden of HF separately, whereas traditional statistical methods are unable to reduce the collinearity between these factors.^21–23^ Age effect refers to the effect of changes in biologic aging. Period effect represents the influence of human factors on HF burden that shared by all age groups simultaneously, such as new equipment and medication. Cohort effect reflects changes in HF burden among people of different birth years, which are influenced, for example, by exposure to risk factors. We employed 5 pivotal parameters for the analysis of APC model: net drift, local drift, longitudinal age curve, period rate ratio, and cohort rate ratio. The net drift represents the overall estimated annual percentage change in the ASRs, while the local drift reflects this change over time specific to a certain age group. The longitudinal age curve is the fitted longitudinal age-specific rate in the reference cohort adjusted for period effect. The period rate ratio measures the relative change in HF burden between two distinct time periods, and the cohort rate ratio compares HF burden across groups born in different years. The calculation of these parameters has been detailed in previous study.^20, 22^ To fit the APC model, age groups were defined as 0-4 (<5), 5-9, 10-14, …, and 90-94. The periods were similarly categorized into 5-year intervals (1992-1996, 1997-2001, …, and 2017-2021). Data for the period 1990 to 1991 and age groups >95 years were excluded from the analysis due to their failure to span a 5-year interval.

The autoregressive integrated moving average (ARIMA) model has been introduced to account for the complexity of time series data and is widely used to investigate time-dependent processes in population health.^24^ The basic structure of an ARIMA model is (*p*, *d*, *q*), where *p* is the order of the autoregressive component, *d* is the degree of differencing (integration), and *q* is the order of the moving average component.^25^ To predict future trends of HF over the next 10 years, we applied “auto.arima()” function, an automated algorithm from the “forecast” packages, which simplifies model selection by identifying the best-fitting ARIMA model based on the Akaike Information Criterion (AIC).^26^ Subsequently, the normality and autocorrelation (white noise) of the residuals from the fitted model were evaluated using the residual time series plot, autocorrelation function (ACF) Plot, histogram with density plot, and the Ljung-Box test. A *P*-value greater than 0.05 in the Ljung-Box test indicated no significant autocorrelation, supporting the model’s adequacy.^25^

The joinpoint analysis was performed in joinpoint software (Version 5.2.0). Other analyses and visualizations were conducted in R software (version 4.3.3), utilizing the “ggplot2”, “nih.joinpoint”, “Epi”, “forecast”, and “forest” packages, among others.

### Ethics statement

For GBD studies, the Institutional Review Board of the University of Washington reviewed and approved a waiver of informed consent (https://www.healthdata.org/research-analysis/gbd).

## Results

### Temporal trends in the burden of HF

In 2021, there were 13099727 (95% UI, 11320895 to 15376467) HF cases in China, with ASRs of 692.50 (95% UI, 607.26 to 802.00) per 100000 population. This represents an increase from 1990, when there were 4683303 (95% UI, 4036499 to 5435212) cases and ASRs of 644.58 (95% UI, 558.65 to 751.14) per 100000 population. Additionally, the number of YLDs more than doubled from 459512 (95% UI, 313552 to 630785) in 1990 to 1290810 (95% UI, 865894 to 1775731) in 2021, while the ASRs increased at AAPC of 0.25% (95% CI, 0.23 to 0.27) over the same period. **Table 1** presents the prevalence and YLDs in 1990 and 2021, as well as the AAPC from 1990 to 2021, for China and globally. Overall, the burden of HF has been on an upward trend over the last 30 years, with a more pronounced increase in China compared to the global level, as reflected by higher AAPC. Since 1990, the number of HF cases in China has increased by 2.8 times, making up 23.6% of the global total in 2021, compared to a 2.2-fold increase globally. However, from the perspective of ASRs of prevalence and YLDs, HF burden in China is higher than the global average. Trends in HF burden for China and globally over the past 30 years are presented in **Supplemental Figure S1**.

**Table 1.**
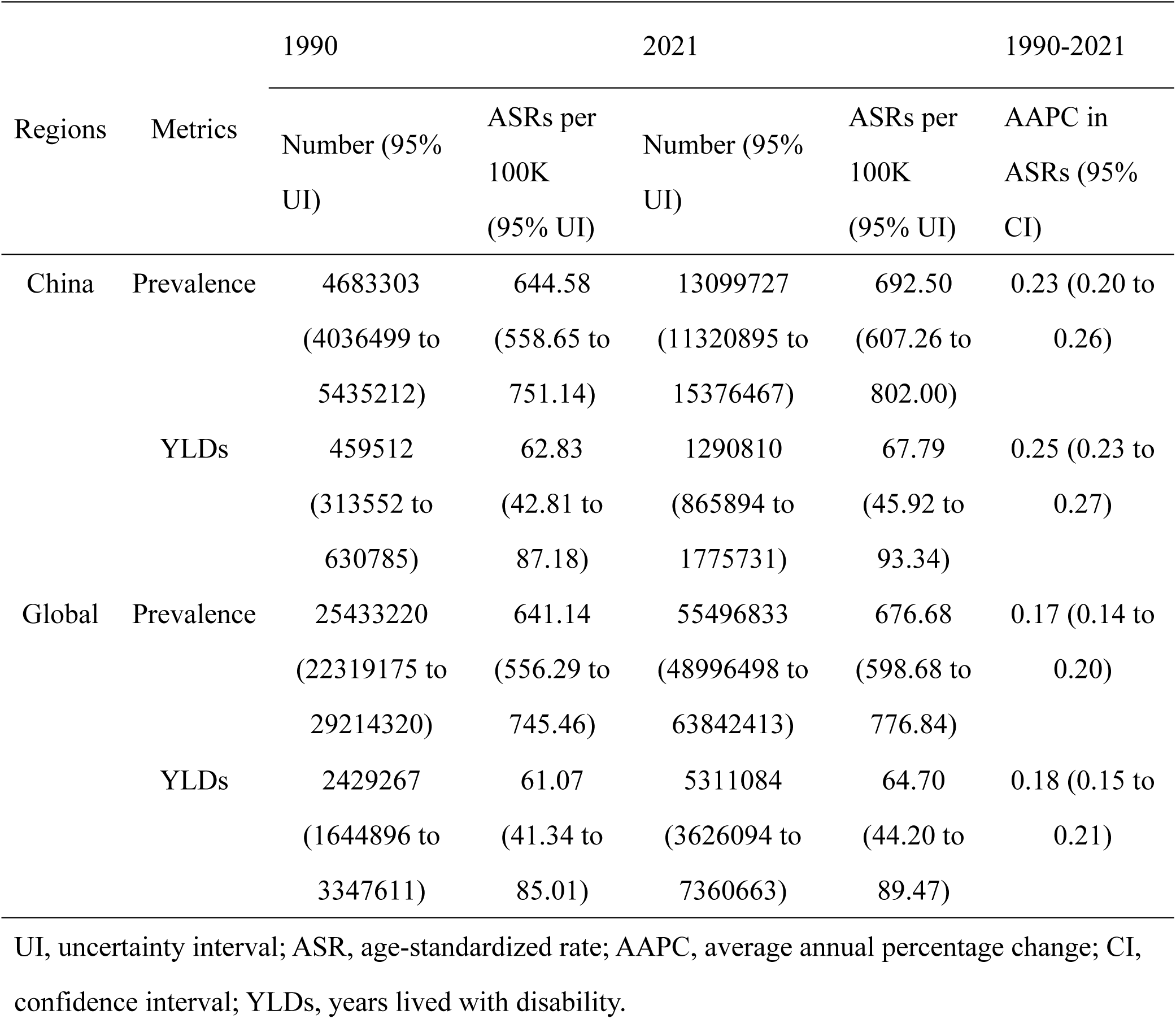
The burden of heart failure in China and globally.

| Regions | Metrics | 1990 |  | 2021 |  | 1990-2021 |
| --- | --- | --- | --- | --- | --- | --- |
|  |  | Number (95%<br>UI) | ASRs per<br>100K<br>(95% UI) | Number (95%<br>UI) | ASRs per<br>100K<br>(95% UI) | AAPC in<br>ASRs (95%<br>CI) |
| China | Prevalence | 4683303 | 644.58 | 13099727 | 692.50 | 0.23 (0.20 to<br>0.26) |
|  |  | (4036499 to<br>5435212) | (558.65 to<br>751.14) | (11320895 to<br>15376467) | (607.26 to<br>802.00) |  |
|  | YLDs | 459512 | 62.83 | 1290810 | 67.79 | 0.25 (0.23 to<br>0.27) |
|  |  | (313552 to<br>630785) | (42.81 to<br>87.18) | (865894 to<br>1775731) | (45.92 to<br>93.34) |  |
| Global | Prevalence | 25433220 | 641.14 | 55496833 | 676.68 | 0.17 (0.14 to<br>0.20) |
|  |  | (22319175 to<br>29214320) | (556.29 to<br>745.46) | (48996498 to<br>63842413) | (598.68 to<br>776.84) |  |
|  | YLDs | 2429267 | 61.07 | 5311084 | 64.70 | 0.18 (0.15 to<br>0.21) |
|  |  | (1644896 to<br>3347611) | (41.34 to<br>85.01) | (3626094 to<br>7360663) | (44.20 to<br>89.47) |  |

UI, uncertainty interval; ASR, age-standardized rate; AAPC, average annual percentage change; CI, confidence interval; YLDs, years lived with disability.

Figure 1 illustrates the trends in sex-specific prevalence and YLDs in China, including both the number and ASRs from 1990 to 2021. The prevalence of HF in both males and females has shown an overall increasing trend, with males consistently experiencing a higher burden than females. Notably, the gap between the cases of HF in males and females gradually widened after 1990 (2446913 vs 2236390), peaking in 2014 (5623587 vs 4598155), when the growth of the prevalence in males slowed, leading to a gradual reduction in the gap, until 2020 (6577128 vs 6075682). The trends in YLDs followed a similar pattern to those of prevalence. These localized trend changes are further described in the results of joinpoint analysis.

**Figure 1.**
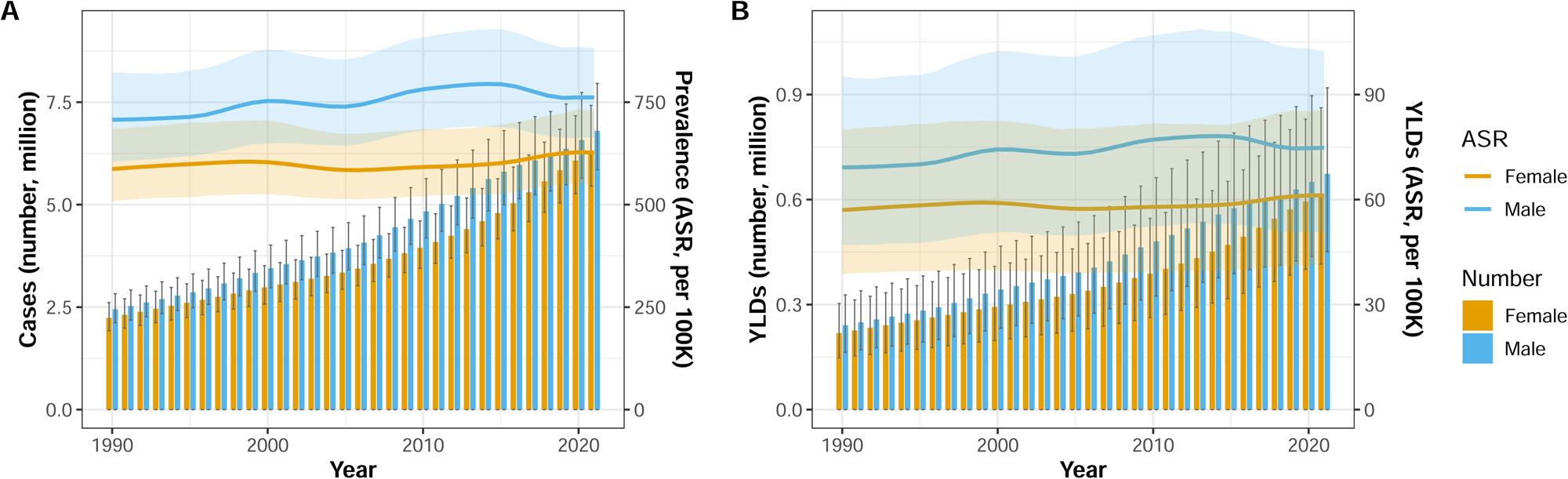
Trends in the burden of heart failure in China by sex from 1990 to 2021. (A) Number of prevalent cases and ASRs. (B) Number of YLDs and ASRs. The translucent ribbons and black error bars represent the 95% UI. ASR, age-standardized rate; YLDs, years lived with disability.

Figure 2 presents the age-specific burden of HF among males and females in China in 2021. Generally, the number of HF cases for both males and females reaches its lowest point in the 20-24 age group (40317 and 31944), then increases with age, particularly rising sharply after the age of 65, and peaking in the 70-74 age group (1372223 and 1121531). Notably, after the age of 80, the number of cases in females surpasses that in males. Similar tendencies are observed in YLDs, where the burden also increases with age and follows a comparable pattern across age groups.

**Figure 2.**
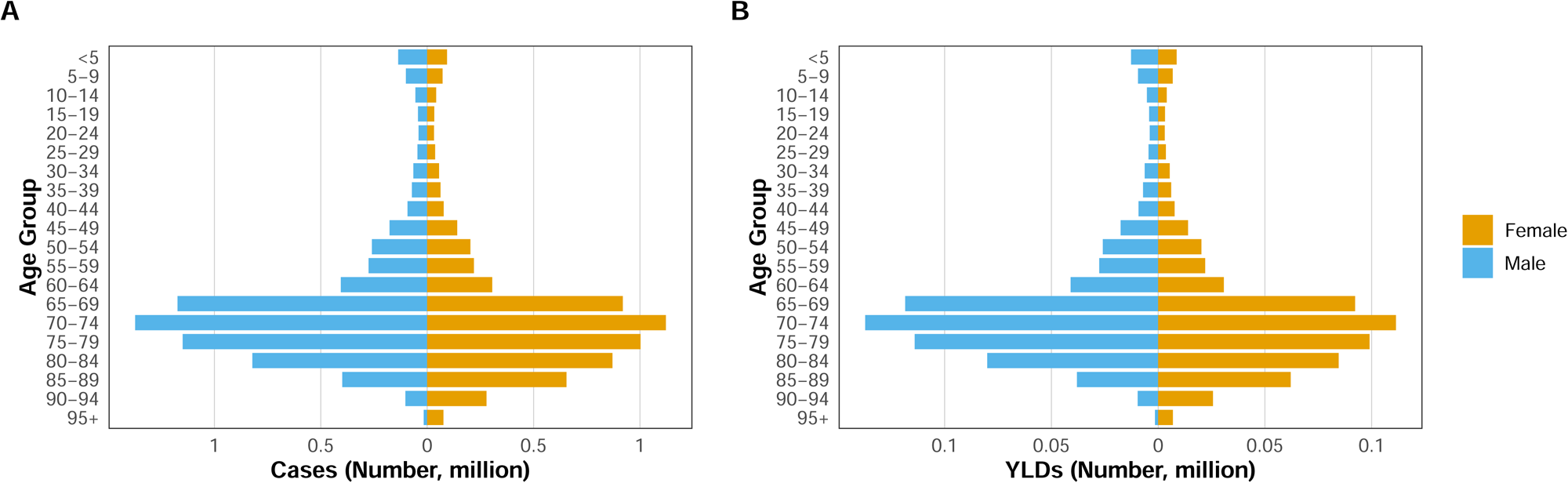
Age-specific burden of heart failure in China, 2021. (A) Number of prevalent cases by age. (B) Number of YLDs by age. YLDs, years lived with disability.

### Joinpoint analysis

The joinpoint analysis clearly quantifies the localized trend changes in ASRs of prevalence and YLDs of HF (Figure 3). It is evident that the burden of HF has shown an upward trend over the past 30 years. After a significant decline in the ASRs of prevalence between 2000 and 2005 (APC = -0.64%), a slow but nonsignificant decline was observed after 2014 (APC = -0.02%). Both males (APC = -0.49%) and females (APC = -0.72%) experienced a significant decline in ASRs of prevalence between 2000 and 2005, while the ASRs of prevalence in males showed another significant decline after 2014 (APC = -0.74%), narrowing the gap in the burden of HF between males and females. Additionally, the ASRs of prevalence in males (AAPC = 0.24%) increased at a faster rate than in females (AAPC = 0.21%). The trend in ASRs of YLDs closely mirrors that of prevalence. Meanwhile, it is noteworthy that the AAPC for the ASRs of YLDs in males, females, and both sexes are higher than that of prevalence. Detailed results of the joinpoint analysis can be found in **Supplemental Table S2**.

**Figure 3.**
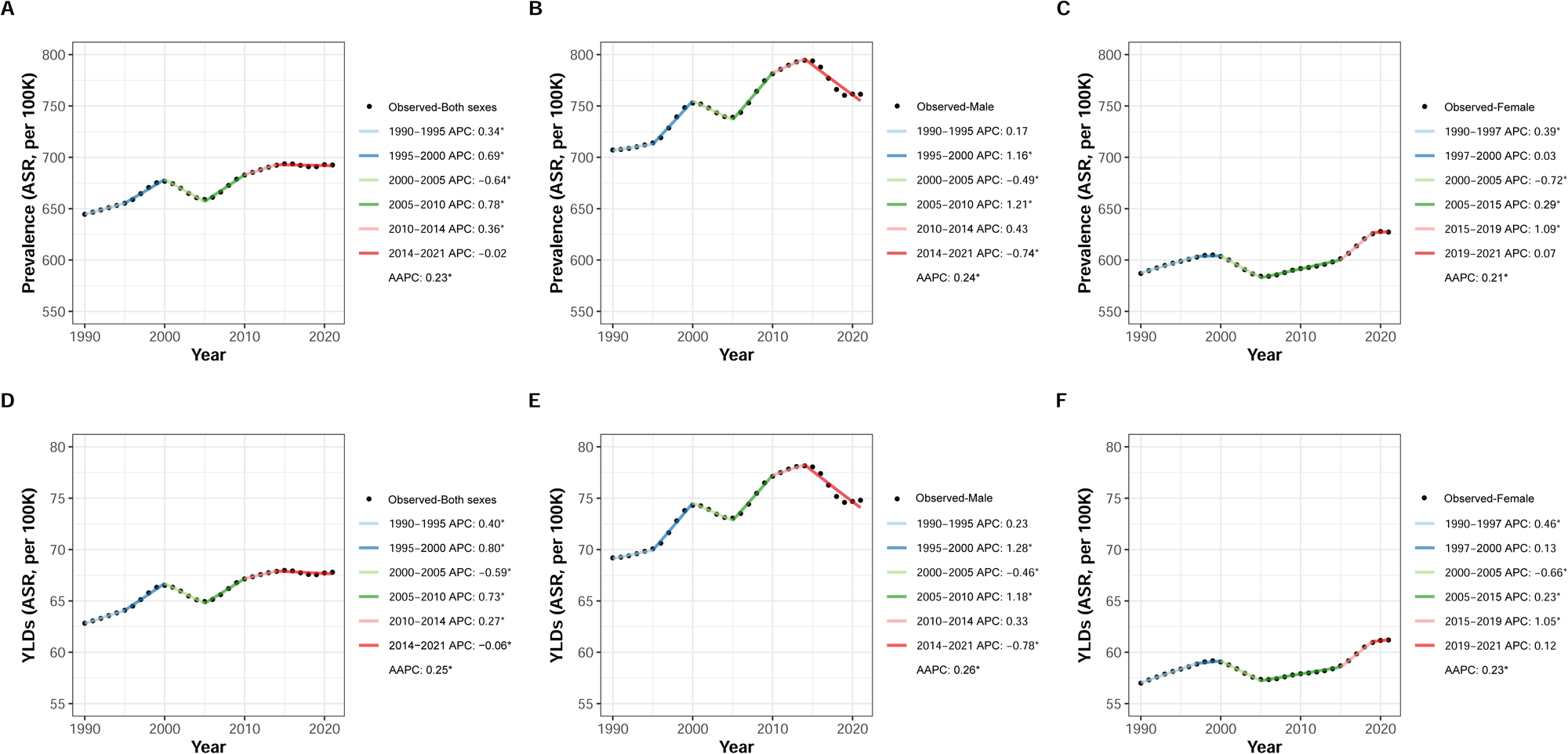
Joinpoint analysis of the burden of heart failure in China from 1990 to 2021. (A-C) Trends in age-standardized prevalence for both sexes, males, and females, respectively. (D-F) Trends in age-standardized YLDs for both sexes, males, and females, respectively. Significant trends are indicated with an asterisk (*) based on *P*-values of APC and AAPC. ASR, age-standardized rate; APC, annual percentage changes; AAPC, average annual percentage change; YLDs, years lived with disability.

### Age-period-cohort effects

The local drift and net drift of HF prevalence calculated using the APC model are shown in Figure 4A. The solid horizontal line and the dashed line represent the net drift (0.97%) with its 95%CI (0.76% to 1.19%), indicating that the overall ASRs of HF prevalence in China continues to increase. The blue curve represents the local drift, which shows an inverted U-shape and intersects with the net drift at one point. The growth rate for younger patients (≤50-54 age group) is higher than the overall level, indicating that the proportion of younger patients within the total HF population has gradually increased from 1992 to 2021. In contrast, the proportion of elderly patients (≥55-59 age group) has shown a downward trend. Additionally, the local drift for patients aged ≥70 is below 0, suggesting a continuous decline in their prevalence from 1992 to 2021.

**Figure 4.**
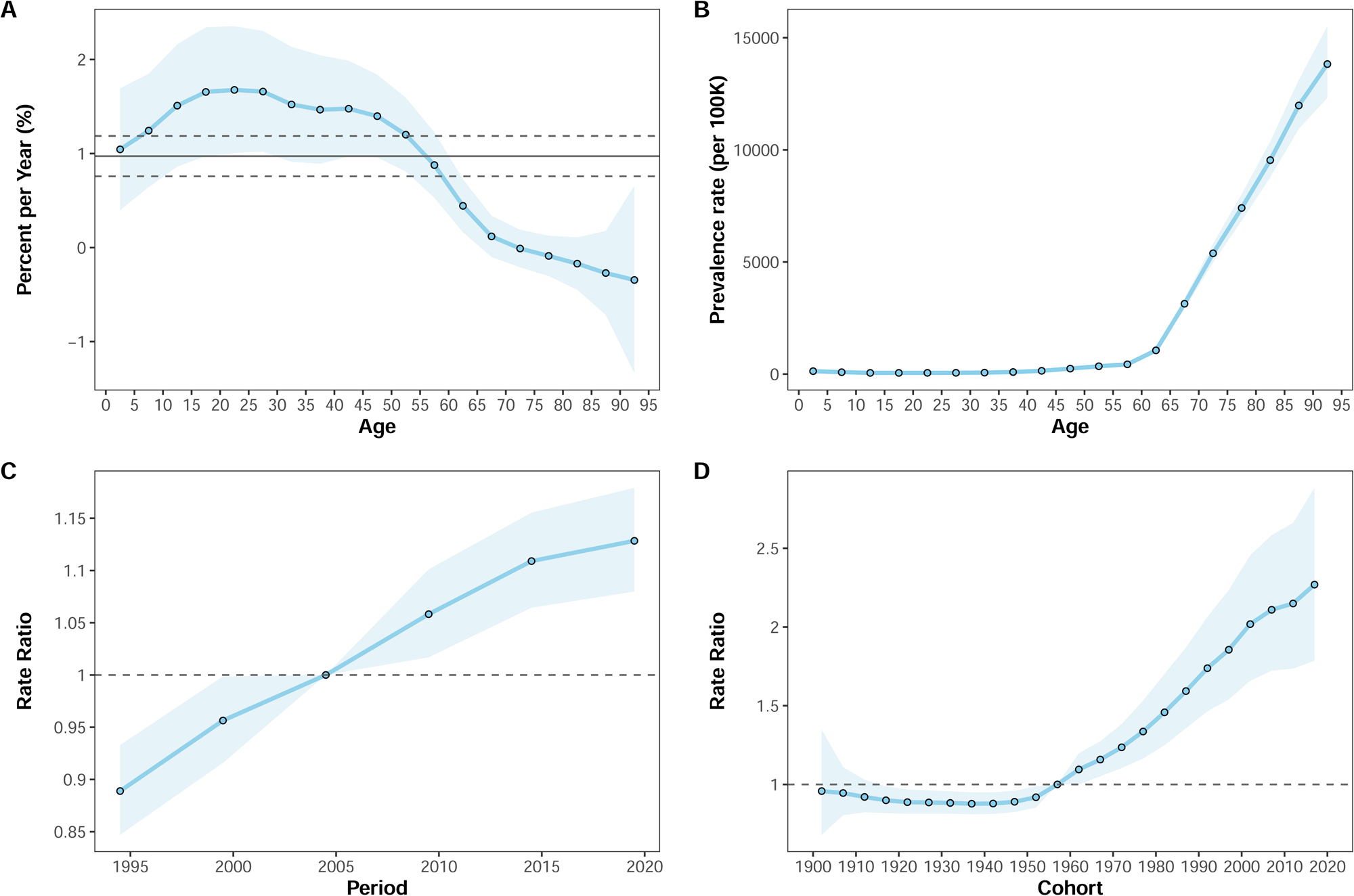
Age-period-cohort effects on the prevalence of heart failure in China from 1992 to 2021. (A) Net drift and local drift for the prevalence of heart failure. The dashed lines and translucent ribbons represent the 95% CI of the net drift and the 95% CI of the local drift, respectively. (B) Longitudinal age curves for the prevalence of heart failure, adjusted for period effect. The translucent ribbons indicate the 95% CI of the age-specific prevalence rate. (C) Period rate ratio of the prevalence of heart failure, adjusted for age and cohort effects. The translucent ribbons indicate the 95% CI of the period rate ratio, and the period 2002 to 2006 is the reference period. (D) Cohort rate ratio of the prevalence of heart failure, adjusted for age and period effects. The translucent ribbons indicate the 95% CI of the cohort rate ratio, and cohort 1957 is the reference cohort.

Figure 4B shows the longitudinal age curve of HF prevalence in China, independent of period effect. The prevalence rate of HF remains stable up to the 60-64 age group, after which it increases rapidly with age. Figure 4C and Figure 4D present the period rate ratio and cohort rate ratio for the prevalence of HF in China. From 1992 to 2021, the period rate ratio for HF showed continuous growth, with the growth rate slowing after the period 2012-2016. The cohort rate ratio for HF showed a slight decline from the 1902 to 1937 birth cohorts, followed by a continuous increase in more recent birth cohorts. The age-period-cohort effects on YLDs are very similar to those on prevalence, and thus will not be elaborated here (**Supplemental Figure S2**).

### Trends in causes of HF

Given that the majority of the HF burden is attributable to a few major causes, we selected the top 10 contributors in 2021 and plotted their trends from 1990 to 2021 (Figure 5). Focusing on these top contributors allows us to gain clearer insights into the primary drivers of disease burden. It is evident that the prevalence and YLDs of HF caused by ischemic heart disease and hypertensive heart disease far exceed those caused by other conditions. The contributions of ischemic heart disease, congenital birth defects, and cardiomyopathy and myocarditis to the prevalence of HF have steadily increased, and their rankings have risen compared to 1990. Notably, the most significant increase was observed in ischemic heart disease, which surpassed hypertensive heart disease after 2002 to become the leading cause of HF. Although the prevalence of HF attributable to stroke is lower than that attributable to COPD, the YLDs attributable to stroke surpassed those of COPD after 2000.

**Figure 5.**
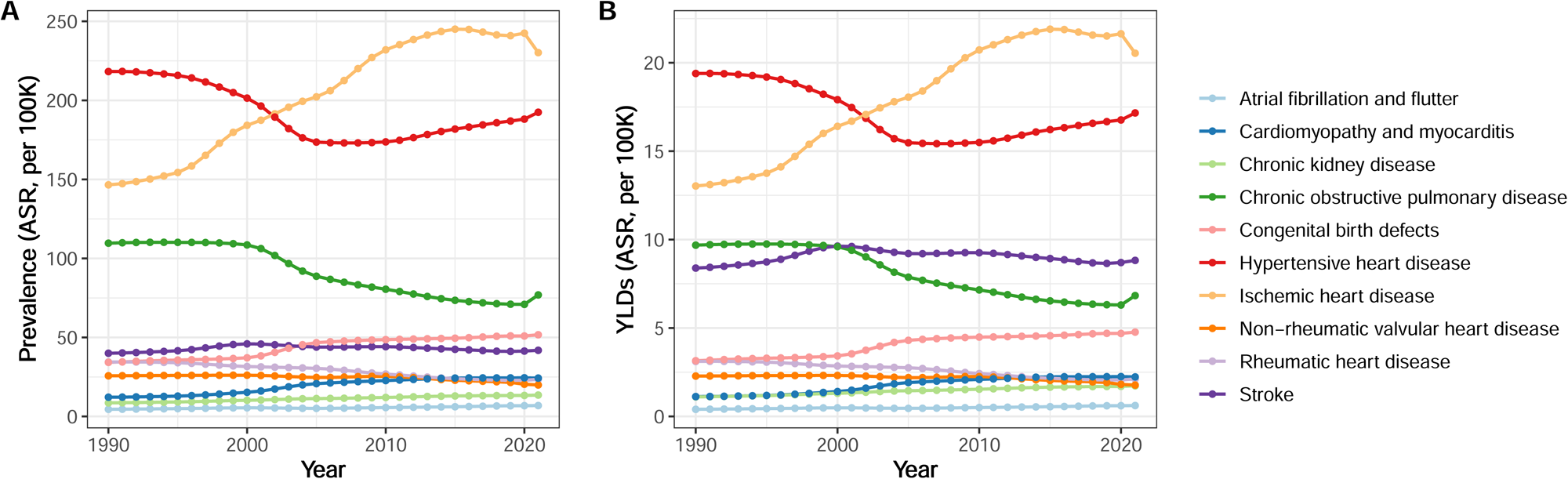
Trends in age-standardized rates of prevalence (A) and YLDs (B) attributable to major causes of heart failure in China from 1990 to 2021. ASR, age-standardized rate; YLDs, years lived with disability.

Furthermore, a Sankey diagram is used to depict the proportions of sex, cause, and age distribution of HF in 2021, offering a clear overview of how these factors contribute to HF burden (**Supplemental Figure S3**). Among all patients, the elderly (≥55 years) made up the largest proportion. Ischemic heart disease and hypertensive heart disease were the leading causes of HF in both the middle-aged (20-54 years) and elderly (≥55 years) patients, whereas congenital birth defects were the primary cause in younger patients (<20 years).

### Prediction of the HF burden in 2022–2031

The ARIMA model is used to predict the trends in the prevalence and YLDs of HF over the next 10 years (Figure 6). The best-fitting model for the ASRs of prevalence is identified as (2, 1, 1), with an AIC value of 93.15. The residual time series plot, ACF plot, histogram with density plot, and the Ljung-Box test (χ² = 0.095, P = 0.757) confirm the normality of residuals and lack of autocorrelation, suggesting the model’s adequacy (**Supplemental Figure S4**). Using the same approach, the ARIMA model (2, 1, 0) for the ASRs of YLDs (AIC = -53.68) is developed. The adequacy of the model is validated by the aforementioned plots (**Supplemental Figure S5**) and the Ljung-Box test (χ² = 1.630, P = 0.202).

**Figure 6.**
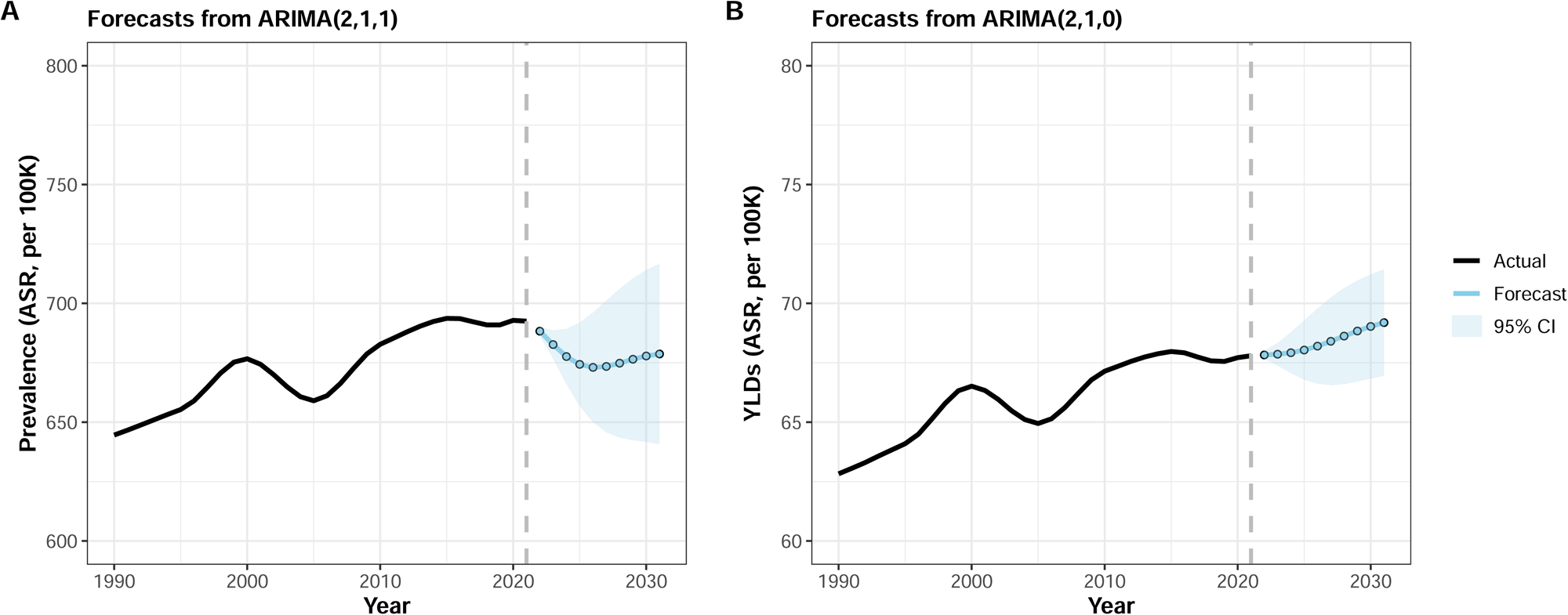
Predicted trends in age-standardized rates of heart failure prevalence (A) and YLDs (B) in China from 2022 to 2031. The black lines represent the trend over the past 30 years, while the blue lines with black dots represent the forecast for the next 10 years. ARIMA, the autoregressive integrated moving average; ASR, age-standardized rate; YLDs, years lived with disability.

Based on the fitted models, the ASRs of prevalence are projected to decrease from 688.28 (95% CI, 686.44 to 690.12) in 2022 to 673.05 (95% CI, 649.94 to 696.16) in 2026, followed by a slow increase, reaching 678.69 (95% CI, 640.75 to 716.63) in 2031. In contrast, the ASRs of YLDs are expected to continue increasing, from 67.83 (95% CI, 67.65 to 68.00) in 2022 to 69.19 (95% CI, 66.95 to 71.43) in 2031.

## Discussion

To our knowledge, this is the first analysis of the GBD 2021 that comprehensively describes the burden of HF in China. Over the past 30 years, both the burden of HF and its growth rate in China have exceeded the global levels. In 2021, approximately 13.1 million individuals lived with HF and this illness accounted for 1.3 million YLDs in China. This is generally consistent with the prevalence estimated using the China Hypertension Survey and the National Urban Employee Basic Medical Insurance Database.^27, 28^ Specifically, the burden is higher in males than in females, and it is the heaviest among the elderly. As for the causes of HF, ischemic heart disease and hypertensive heart disease stand out as the most striking causes among all patients, while congenital birth defects are the leading cause among children and adolescents.

The algorithms and methodologies of the GBD study are continuously being optimized, making it essential to utilize the most up-to-date data.^17^ Our analysis of GBD 2021 reveals that both ASRs of prevalence and YLDs in China and globally have increased in 2021 compared to 1990. This trend is significantly different from what was presented in GBD 2019.^12^ The joinpoint regression model indicates that ASRs of prevalence and YLDs have ceased to increase after 2014, which may reflect the positive outcomes of China’s continuous improvements in HF management.^29, 30^ This stabilization is largely attributed to the significant reduction in the burden of HF among males (Figure 3B**, 3E)**.

Males generally bear a heavier burden than females in terms of risk factors and causes of HF,^31, 32^ which aligns with the observed gender differences in the disease burden. Therefore, it is no surprise that, under the same management, the improvement in HF burden is more pronounced in males.

Although ASRs of prevalence and YLDs have stabilized, HF remains a huge health care burden in China due to population aging and the increasing absolute number of patients.^28, 33^

The results of APC effects have further revealed the severe nature of the HF burden in China. The longitudinal age curve shows a positive correlation between age and HF burden, consistent with previous studies indicating that age is a risk factor for HF.^34, 35^ The prevalence and YLDs rates of HF increase rapidly after the 60-64 age group, suggesting that greater efforts are needed to monitor and prevent HF in individuals aged 65 and above.

The curve of local drift suggests that the proportion of younger patients among the HF population is gradually rising. In contrast, HF burden in individuals aged ≥55-59 is growing at a rate slower than the overall average, even the burden among those aged ≥70 is declining. Several factors may explain this trend. First, elderly patients are consistently a primary focus and may benefit more from the prevention and management efforts. Second, the unfavorable mortality rates^36^ and survival rates^37^ of HF may lead to the premature death of some patients, while those who survive tend to be relatively healthier.

Since the 2009 health-care reform, the Chinese government has doubled its expenditure in the health sector and established an integrated health-care delivery system based on primary health care.^38, 39^ However, according to the trend of the period effect, the risk of HF continues to increase, which is closely associated with changes in certain lifestyles and risk factors.

Diet, as one of the most important components of the Chinese lifestyles, has undergone significant changes over the past few decades. People have shifted from a predominantly plant-based diet to a Western-style diet characterized by high fat and high animal-based food intake.^40^ The intake of energy, carbohydrates, and protein has decreased, whereas fat consumption has significantly increased.^40^ Furthermore, the consumption of whole grains, vegetables, and fruits in China over the past 20 years has remained far below the levels recommended by national dietary guidelines, and their inadequate intake constitutes a major dietary risk factor for ischemic heart disease and stroke.^41^ Moreover, sodium intake in China has decreased since 1991, but it remains significantly higher than the World Health Organization (WHO) recommended level of <2.0 g/day, which is a major risk factor for hypertension.^42^ The harmful effects of tobacco on cardiovascular and pulmonary health are well-documented. As the largest tobacco-consuming country, China currently has over 300 million smokers, with approximately half of them being tobacco dependent, making it more difficult for them to initiate or succeed in quitting smoking.^43^ Unfortunately, younger age and lower socioeconomic status are associated with a higher risk of tobacco dependence.^43^ Physical activity has been proven to be associated with a lower risk of cardiovascular disease (CVD).^44^ Over the past decade, the prevalence and volume of physical activity among adults have increased slightly, but they still remain at low levels. In contrast, the prevalence and volume of physical activity among children and adolescents have shown a declining trend.^45, 46^

Obesity, dyslipidemia, hypertension, and diabetes are fundamental cardiovascular risk factors, and their worsening trends also explain the continuous increase in HF risk. Data from the China Chronic Disease and Risk Factors Surveillance program indicate that the standardized mean body mass index (BMI) levels rose from 22.7 kg/m² to 24.4 kg/m², and the prevalence of obesity increased from 3.1% to 8.1% over the past decade.^47^ Another study comparing three national cross-sectional surveys found that the prevalence of dyslipidemia among Chinese adults increased from 2002 to 2015, and the weighted mean of low-density lipoprotein cholesterol (LDL-C) increased from 2.12 mmol/L to 2.87 mmol/L.^48^ Multiple nationwide cross-sectional studies indicated that the prevalence of hypertension increased from 5.1% in 1958 to 27.5% in 2018, with awareness and treatment rates below 50%.^15^ Meanwhile, the prevalence of diabetes rose from 0.67% in 1980 to 11.2% in 2017.^15^

To our relief, after the period 2012-2016, the growth of period rate ratio began to slow. This improvement can be attributed not only to sustained financial support but also to significant progress in the quality of HF management in China, including standardized diagnosis, guideline-directed medical therapies, and device therapies.^49^ The trend of the period effect is likely to improve further with a deeper understanding of HF and the establishment of the four pillars of HF therapy after 2021.^50, 51^ While the period effect has a broad impact across all age groups, the cohort effect highlights that younger generations are at higher risk of HF. Individuals from more recent birth cohorts are exposed to Westernized diets and other unhealthy lifestyles at a younger age, and they are also more vulnerable to risk factors of HF during their growth. In this context, it is essential to strengthen health education and regular screening for children and adolescents to mitigate the risk of early onset HF.

In line with global trends,^13^ hypertensive heart disease and ischemic heart disease are the leading causes of HF in China. To address these major causes, we believe it is crucial to enhance public health education to improve awareness and treatment rates, while general practitioners need to identify and manage related risk factors as early as possible. This presents a challenge to China’s primary health care system. For children and adolescents, congenital heart disease ranks first among congenital birth defects, which is the main cause of HF.^15^ A meta-analysis has filled the gap left by the GBD study, showing that atrial septal defect, patent ductus arteriosus, and ventricular septal defect are the most common subtypes of congenital heart disease in China, accounting for 27.37%, 20.66%, and 18.72%, respectively.^52^ Since the establishment of a screening and diagnosis system for congenital heart disease by the Chinese government in 2002, prenatal screening coverage and detection rates have continuously improved, which partly explain the slight increase in congenital birth defects after 2000.^53^ However, China still faces issues regarding insufficient screening, diagnosis, and treatment capabilities in central and western regions, as well as rural areas.^53^ Besides, the burden of HF attributable to COPD has declined significantly after 2000, largely due to the strengthened anti-smoking policies.^54, 55^ It is noteworthy that ASRs of prevalence attributable to stroke rank fifth, whereas ASRs of YLDs rank third, indicating that HF caused by stroke has a pronounced impact on patients’ quality of life. Considering that stroke and HF share several major risk factors,^56^ clinicians must pay close attention to the cardiac function of stroke patients when initiating therapies. Early identification and intervention for HF are important to reduce the impact on the quality of life.

The course of HF is prolonged, and symptoms often lag behind the progression of the disease itself. The ARIMA model predicts that ASRs of prevalence will first decrease and then slightly increase over the next decade. This trend may be the result of continuous improvements in HF management, sustained financial support, and the high mortality rates among elderly patients. On the other hand, the continuous increase in ASRs of YLDs suggests that the quality of life of HF patients will further deteriorate. This is associated with worsening HF risk factors and its earlier onset tendency, as the increasing proportion of younger patients indicates that more individuals will be affected by HF for longer durations. The results of ARIMA model indicate that improving patients’ quality of life will be a prominent challenge in the future. We believe that, in addition to pharmacological treatments, cardiac rehabilitation is a promising solution. Cardiac rehabilitation is a comprehensive intervention that includes, but is not limited to, exercise training, nutritional advice, patient education, risk factor control, and psychological support.^57^ High quality studies have shown that cardiac rehabilitation can effectively reduce the risk of hospitalization in HF patients, while also lowering healthcare costs and improving quality of life.^58^ Cardiac rehabilitation is even considered the fifth pillar in the management of HF.^59^ However, the importance attached to cardiac rehabilitation in China is far from sufficient. A cross-sectional survey of 124 large medical centers in China showed that only 30 of them provided cardiac rehabilitation services, and the estimated availability of cardiac rehabilitation programs was about 2 programs per 100 million inhabitants, which is much lower compared to the availability in the United States.^60, 61^ We recommend increasing awareness of the benefits of cardiac rehabilitation among both patients and medical staff, and actively promoting cardiac rehabilitation programs in hospitals. This is of great value in improving the quality of life and mitigating the growth of YLDs.

We conducted a comprehensive analysis of the changing patterns of HF in China using the latest GBD 2021 data, but several limitations should be considered when interpreting the results. First, we were unable to report the burden of specific HF subtypes (HF with reduced/mildly reduced/preserved ejection fraction, HFrEF/HFmrEF/HFpEF). Since each type differs in epidemiology, pathophysiology, treatment, and prognosis,^27, 62–65^ this prevents us from providing more tailored advice on healthcare strategies. Second, there are disparities in economic development and healthcare quality across provinces in China, yet we were unable to obtain provincial-level data to describe the geographical distribution of HF burden. Third, in the GBD study, HF is categorized as an “impairment”, which limits its burden assessment to prevalence and YLDs. The lack of data on HF mortality and YLLs may hinders the accurate description of its threat to public health, which further impacts the allocation of healthcare resources and policymaking. Interestingly, CKD, which is also an end-stage disease with a similar five-year survival rate to HF,^37, 66^ is categorized as a “cause of death or injury”. The classification of HF in the GBD study may need reconsideration in the future. Lastly, the ARIMA model is a time series model which integrates the comprehensive effects of influencing factors into the time variable, characterized by simplicity and efficiency.^67^ However, relying solely on the time variable makes it difficult to predict the long-term trend of HF burden.

Particularly, when new influencing factors emerge in the future, such as advances in treatment or policy changes, the predictive power may decline. Therefore, our predictions should be interpreted with caution.

## Conclusions

Although the ASRs of prevalence and YLDs of HF in China gradually stabilized from 1990 to 2021, the burden remains substantial. The HF burden is heavier in males and the elderly, with ischemic heart disease and hypertensive heart disease as the primary causes, while congenital birth defects are the leading causes among children and adolescents. Lifestyle shifts and the worsening burden of cardiovascular risk factors continue to drive the risk of HF, and younger generations tend to be more vulnerable to HF. Given the severity of the HF burden in China, comprehensive strategies must be considered: (1) strengthening the primary healthcare system to improve prevention and early treatment of the major causes (ischemic heart disease, hypertensive heart disease, and congenital birth defects) and common risk factors (obesity, dyslipidemia, hypertension, and diabetes) of HF; (2) enhancing public awareness to encourage healthy lifestyle (balanced diet, reduced salt intake, smoking cessation, and increased physical activity), as well as improving awareness and treatment rates for the disease; (3) promoting cardiac rehabilitation to enhance quality of life and mitigate the growth of YLDs.

## Data Availability

Datasets analyzed for this study are publicly available at https://vizhub.healthdata.org/gbd-results/.

## Non-standard Abbreviations and Acronyms

AAPC: Average Annual Percentage Changes
ACF: Autocorrelation Function
AIC: Akaike Information Criterion
APC: Annual Percentage Changes
The APC model: The age-period-cohort model
ARIMA: Autoregressive Integrated Moving Average
ASR: Age-Standardized Rate
DALYs: Disability-Adjusted Life-Years
GBD: Global Burden of Disease
HAQ: Healthcare Quality and Access
HF: Heart Failure
MR-BRT: Meta-Regression—Bayesian, Regularized, Trimmed
UI: Uncertainty Interval
WHO: World Health Organization
YLDs: Years Lived with Disability
YLLs: Years of Life Lost

## Acknowledgments

The authors thank the GBD 2021 Diseases and Injuries Collaborators and the Institute for Health Metrics and Evaluation (IHME) for sharing the high-quality data.

